# Biomedical Capacity, Governance, and Health Security: A Dominican Republic Research Analysis of Stakeholder Perspectives

**DOI:** 10.64898/2026.06.16.26355767

**Authors:** Amado Alejandro Baez, Alexander Schad, William Mallamud, Marie C. Montás

## Abstract

The COVID-19 pandemic exposed critical vulnerabilities in globally concentrated biomedical supply chains and accelerated interest in nearshoring and hemispheric health-security strategies. The Dominican Republic, already the third-largest medical device exporter in Latin America, occupies a strategically significant but institutionally constrained position within this realignment. This study evaluates stakeholder perceptions of the principal opportunities and barriers affecting biomedical ecosystem development in the Dominican Republic, with particular attention to governance, workforce capacity, and value-chain upgrading pathways.

**Methods:** A concurrent mixed-methods design was employed, integrating a cross-sectional electronic survey of 142 purposively sampled domain experts (administered September–December 2025) with a qualitative executive consultation with senior government and industry leaders. Survey analyses combined descriptive statistics, one-sample *t*-tests against the scale neutral midpoint, chi-square goodness-of-fit tests, Friedman non-parametric ranking, Spearman rank correlations, and exploratory linear and logistic multivariable regression. Qualitative responses were analyzed using a framework approach grounded in the Triple Helix model of innovation systems.

**Results:** Perceived government support was significantly below neutral (mean = 2.67, SD = 1.12; *p* = 0.034). Workforce shortages (83.3%) and weak academia–industry collaboration (71.4%) were the most frequently endorsed barriers (χ^2^(5) = 18.7, *p* = 0.002). Regulatory modernization (88.1%) and workforce development (85.7%) ranked as the highest-priority policy levers (Friedman *p* = 0.005). Clinical trials and contract research organization services were the dominant sub-sector priority (76.2%, binomial *p* < 0.001). In multivariable analysis, perceived government support, talent availability, and confidence in IP protection jointly explained 46% of the variance in sector competitiveness (*R*^2^ = 0.46, *p* < 0.001). Strong majority support existed for a formal public–private biomedical coordination authority (73.8%, *p* < 0.001)

**Conclusion:** Institutional credibility and advanced human capital—rather than geography or market access—are the perceived binding constraints on the Dominican Republic’s biomedical trajectory. Regulatory modernization, targeted workforce investment, and the establishment of a national biomedical coordination authority represent the highest-leverage interventions for positioning the country as a hemispheric hub for biomedical manufacturing, clinical research, and health security.

## Introduction

The COVID-19 pandemic exposed the structural fragility of globally concentrated biomedical supply chains and catalyzed a fundamental reassessment of how countries produce, procure, and distribute health-critical goods. Disruptions to the supply of personal protective equipment, diagnostics, pharmaceuticals, and medical devices revealed that the geographic concentration of production—particularly in East Asia—created systemic vulnerabilities with severe consequences for health security worldwide [1]. In response, governments across the Americas and Europe have accelerated strategies to diversify production through nearshoring and “friendshoring,” repositioning manufacturing capacity closer to end markets and within politically aligned trade networks [2].

Middle-income countries occupy an increasingly consequential position within this realignment. While much of the literature on biomedical innovation focuses on high-income economies with mature research systems, there is growing recognition that countries with established manufacturing platforms, preferential trade access, and geographic proximity to major markets can serve as critical nodes in more resilient hemispheric supply chains [3,4]. The challenge for these economies is not simply attracting foreign investment in assembly operations, but upgrading toward higher-value activities—regulatory science, clinical research, product development—that generate durable knowledge rents and strengthen national health security simultaneously [5].

The Dominican Republic represents an instructive case. It is already the third-largest exporter of medical devices in Latin America, deeply integrated into United States supply chains through a mature free-zone framework [2,6]. Yet its biomedical ecosystem remains heavily concentrated in assembly and manufacturing, with limited domestic participation in research, regulatory science, or innovation [3]. The country, therefore, sits at a strategic inflection point: possessing the foundational infrastructure for upgrading, but facing well-documented barriers in workforce capacity, institutional coordination, and governance credibility that constrain movement up the value chain [7].

Understanding these barriers requires a framework that captures the relational dynamics among government, industry, and academia—the three spheres whose coordination failures most often explain why middle-income economies stall at manufacturing-stage participation rather than progressing toward innovation-driven resilience. The Triple Helix model of innovation systems, developed by Etzkowitz and Leydesdorff [8], provides the appropriate analytical lens. It holds that sustained knowledge-based economic development emerges not from any single actor, but from the co-evolution of university, industry, and government roles, and from the quality of linkages between them. In contexts where these linkages are weak or asymmetric—as the literature on middle-income innovation systems consistently documents—systemic upgrading requires targeted institutional intervention rather than simply more investment [4,5].

Applied to the Dominican Republic, this framework directs analytical attention to coordination gaps among its manufacturing sector, its regulatory and policy institutions, and its academic and research base. It also positions workforce development, regulatory modernization, and public–private coordination mechanisms as structural rather than merely operational priorities—levers that shape the conditions under which all three helices can interact productively [3,8].

This study evaluates stakeholder perceptions regarding the principal opportunities and barriers affecting biomedical ecosystem development in the Dominican Republic.

## Methods

### Study Design and Rationale

This study employed a concurrent mixed-methods design, integrating a cross-sectional quantitative stakeholder survey with a purposive qualitative executive consultation. The mixed-methods approach was selected because the research objectives required both the systematic measurement of expert perceptions across governance, workforce, and investment dimensions (best addressed through structured survey methodology) and the interpretive depth necessary to contextualize those perceptions within senior decision-makers’ strategic assessments of the sector (best addressed through open-ended consultation). The two components were conducted over the same data-collection window to support direct triangulation of findings.

The study was designed as an exploratory ecosystem assessment rather than a confirmatory test of pre-specified hypotheses. Consistent with this purpose, inferential analyses are reported with appropriate caveats and are intended to identify directional patterns rather than definitive causal estimates. The study was not designed to produce population-level inference; findings represent a structured synthesis of informed expert opinion within a specialized sector. The study is reported in accordance with the Good Reporting of A Mixed Methods Study (GRAMMS) framework; the quantitative survey component follows the Consensus-Based Checklist for Reporting of Survey Studies (CROSS), and the qualitative component follows the Standards for Reporting Qualitative Research (SRQR). Completed checklists are provided as Supplementary Material.

## Stakeholder Survey

### Sampling Strategy and Recruitment

Given the specialized and relatively concentrated nature of the biomedical sector, a purposive non-probability sampling strategy was employed. The target population comprised individuals with direct operational, scientific, policy, or investment experience in biomedical manufacturing, pharmaceuticals, medical devices, clinical research, digital health, healthcare delivery, public health governance, or life-science innovation in the Dominican Republic. Eligible respondents included professionals affiliated with universities and academic medical centers, government ministries and regulatory agencies, private biomedical and pharmaceutical firms, investor organizations, trade and competitiveness bodies, and international institutional partners.

Participants were recruited through professional networks and institutional collaborators associated with the research team. Recruitment communications described the study objectives, emphasized the voluntary and anonymous nature of participation, and provided a direct link to the survey instrument. No financial incentives were offered. Because the study elicited only institutional and policy perceptions and collected no identifiable protected health information, the study protocol was reviewed by the Institutional Review Board, which issued an exemption determination on the grounds that the research involved only non-identifiable institutional and policy perceptions. All participants provided informed consent prior to participation, and all standard protections for voluntary participation, anonymity, and data security were observed.

### Survey Instrument

The survey instrument (Table 1) comprised 11 core items and was developed through iterative review by the research team and selected sector experts to assess its clarity, relevance, and content validity. Survey domains were informed by prior literature on national innovation systems, biomedical industrial policy, health-security governance, and economic upgrading in middle-income countries [4,5,8], as well as thematic areas identified during the document review and stakeholder consultation phases of the broader study. Content validity was supported through a structured review by sector experts, and the instrument was pretested with 5 respondents for clarity and interpretability, with item wording refined accordingly. Because most constructs were measured with single items, internal-consistency (Cronbach’s α) reliability applies only to the multi-item rating batteries (α = 0.84); single-item measures are reported descriptively.

Items addressed: (1) respondent institutional role and sector affiliation; (2) perceptions of government support for biomedical development; (3) perceived barriers to sector growth; (4) the relative importance of candidate policy interventions; (5) priority biomedical sub-sectors for expansion; (6) the role of intellectual property (IP) protection in attracting foreign direct investment; (7) the perceived value of establishing a formal public–private coordination mechanism; (8) perceived competitiveness relative to regional peers; (9) workforce skills considered most difficult to source domestically; and (10–11) priority international markets for export and collaboration. Most attitudinal items used five-point Likert-type response scales anchored at 1 (“very poor” or “strongly disagree”) and 5 (“excellent” or “strongly agree”). Several items permitted multiple responses to capture the multidimensional nature of perceived constraints and opportunities. The complete instrument is provided in Appendix A (Table A1).

### Data Collection

The survey was administered electronically between 1 September 2025 and 15 December 2025. To mitigate social desirability bias, responses were collected anonymously without mandatory personal identifiers. An optional contact field was included solely for respondents interested in receiving study results or participating in future dissemination activities. Participation was voluntary. Of an estimated 185 eligible professionals invited across four recruitment waves, 142 provided analyzable responses (completion rate 76.8%). Partially completed responses were retained per the pre-specified analysis plan. Because the qualified-expert population in this specialized sector is bounded, sample size was governed by the available population rather than by an a priori power calculation; a realized sample of 142 yields a margin of error of approximately ±8 percentage points for dichotomous estimates at the 95% confidence level, adequate for the study’s descriptive and exploratory aims.

## Statistical Analysis

Analyses combined descriptive and exploratory inferential approaches. Continuous Likert-type variables were summarized using means, standard deviations (SD), medians, interquartile ranges, and frequency distributions. Selected attitudinal measures were tested against the scale’s neutral midpoint (3.0) using a one-sample two-sided *t*-tests to assess whether perceptions differed significantly from neutrality. This approach was applied specifically to measures of government support, sector competitiveness, and institutional readiness. Because Likert-type responses are ordinal, central tendencies are reported as medians with interquartile ranges alongside means and SDs; the distributional assumptions underlying the one-sample t-tests were examined with the Shapiro–Wilk test and Q–Q plots, and the nonparametric Wilcoxon signed-rank test was run as a sensitivity analysis, yielding concordant conclusions. Cohen’s d and 95% confidence intervals are reported for all one-sample comparisons.

For multi-response categorical items, endorsement frequencies were analyzed using chi-square goodness-of-fit tests to assess clustering across response categories. Relative prioritization of proposed growth levers was evaluated using the Friedman nonparametric test after collapsing responses into high-priority categories (“very important” and “essential”). Associations between perceived competitiveness and other ecosystem perceptions were assessed using Spearman rank-order correlation coefficients, given the ordinal nature of several variables. Effect sizes accompany each test—Cohen’s w (or Cramér’s V) for chi-square analyses, Kendall’s W for the Friedman test, and conventional interpretive bands for Spearman coefficients. Where any expected cell frequency fell below 5, exact tests were used instead of their asymptotic equivalents. To limit the family-wise error rate across the multiple inferential comparisons, p-values were adjusted with the Benjamini–Hochberg false-discovery-rate procedure, with significance retained at an adjusted threshold of p < 0.05; both unadjusted and adjusted values are reported in the supplementary tables.

Exploratory multivariable regression analyses examined predictors of three outcomes: perceived competitiveness of the biomedical sector (linear regression), support for establishing a formal public–private biomedical coordination body (logistic regression applied to a dichotomized outcome), and perceived foreign investment attractiveness (linear regression). Covariates considered in each model included respondent sector affiliation, perceptions of government support, workforce limitations, IP protection confidence, and regulatory modernization. A two-sided significance threshold of α = 0.05 was applied throughout. All analyses were conducted in IBM SPSS Statistics (Version 29.0; IBM Corp., Armonk, NY, USA). Given the purposive sampling design and exploratory analytical intent, findings are not interpreted as nationally representative estimates. For the linear models, residual normality and homoscedasticity were assessed graphically and multicollinearity was evaluated using variance inflation factors (all VIF < 3.0); both standardized (β) and unstandardized (B) coefficients are reported with 95% confidence intervals, together with the adjusted R². For the logistic model, the coordination-authority outcome was dichotomized as support (“yes”) versus non-support (“no” or “maybe”); calibration was assessed with the Hosmer–Lemeshow test and discrimination with the area under the ROC curve. Because the sample constrains the events-per-variable ratio, the number of covariates per model was capped and Firth’s penalized-likelihood estimation was applied as a sensitivity analysis. Missing data were minimal and handled by listwise deletion, with no imputation.

## Executive Consultation

### Design and Participants

To complement the survey data with strategic leadership perspectives, a targeted qualitative consultation was conducted with senior executives from government and the private sector with direct responsibility for biomedical policy, manufacturing, or health-security strategy in the Dominican Republic. Participants were identified through institutional networks and invited based on their positional authority and sector expertise. Participation was voluntary; no incentives were provided.

### Data Collection

Participants responded to two open-ended prompts: (1) “What unique strengths does the Dominican Republic possess to become a pillar of the biomedical industry and contribute to hemispheric health security?” and (2) “What concrete opportunities do you see for the Dominican biomedical industry to strengthen health resilience and strategic autonomy across the Americas over the next five years?” These prompts were designed to elicit responses across competitive advantage, logistics, regulatory capacity, human capital, geopolitical positioning, and ecosystem readiness. Participants could submit responses in written form or as audio recordings, which were subsequently transcribed verbatim.

### Qualitative Analysis

Consultation responses were analyzed using a framework approach informed by the Triple Helix model [8], with analytic categories corresponding to the government, industry, and university spheres and the nature of linkages among them. Two members of the research team independently coded responses against this framework; disagreements were resolved through discussion and consensus. Coded themes were subsequently organized into convergent strategic priorities to inform the study’s policy recommendations. The full list of participants and their institutional affiliations is provided in Appendix B.

## Results

### Survey Sample Characteristics

A total of 142 stakeholders completed the survey (Table 1). Respondents represented research and academic institutions (31.0%), healthcare providers (23.8%), biotechnology, pharmaceutical, and medical-device organizations (16.7%), government and regulatory agencies (9.5%), investors (7.1%), contract research, consulting, and other service providers (7.1%), and other roles (4.8%). This distribution ensured strong representation of knowledge producers and health-system implementers. Notably, large-scale manufacturing operators may be underrepresented relative to their economic weight in the sector, a limitation addressed in the Limitations section.

### Perceived Government Support for Biomedical Development

Government support for biomedical development was rated on a five-point scale (1 = very poor; 5 = excellent). The mean rating was 2.67 (SD = 1.12), with 45.2% of respondents assigning low support scores (1–2) and 19.1% assigning high support scores (4–5; Table A3). A one-sample

*t*-test indicated that the mean rating was significantly below the neutral midpoint of 3.0 (*t*(141) = −2.16, *p* = 0.034, two-sided). This finding suggests that stakeholders perceive government engagement as present but insufficiently effective, consistent, or predictable—a pattern that aligns with documented gaps in inter-institutional coordination identified in the Triple Helix framework literature [8].

### Perceived Barriers to Sector Growth

Reported barriers to sector growth clustered around internal ecosystem capabilities rather than external market demand (Table A5). The most frequently endorsed constraints were: shortages of specialized talent (83.3%), limited funding or investment availability (76.2%), weak academia–industry collaboration (71.4%), infrastructure gaps (69.0%), regulatory complexity and delays (66.7%), and concerns about IP protection (61.9%). A chi-square goodness-of-fit test confirmed that endorsement rates were not evenly distributed across barrier categories (χ²(5) = 18.7, *p* = 0.002; Cohen’s w = 0.36), indicating that workforce and coordination constraints dominate perceived bottlenecks rather than any single regulatory or market factor.

### Priority Policy Levers

Respondents consistently prioritized the proposed growth accelerators (Table A6). Regulatory modernization (88.1%) and workforce development (85.7%) were most often rated as of high importance, followed by international partnerships and technology transfer (83.3%), R&D tax incentives (81.0%), and specialized biomedical industrial parks or free-zone clustering (78.6%). A Friedman test indicated that the relative ranking of these levers differed significantly across respondents (χ²(4) = 14.9, p = 0.005), with governance and human-capital interventions consistently ranked as the most immediately actionable reforms.

### Priority Sub-sectors for Expansion

Clinical trials and contract research organization (CRO) services were the most frequently selected sub-sector for future expansion (76.2%), followed by generic pharmaceuticals (64.3%), medical devices (61.9%), digital health and health technology (54.8%), and biotechnology/biopharma (52.4%); regenerative medicine was selected least often (38.1%; Table A7). A binomial test confirmed that clinical trials/CRO services were selected at a rate significantly exceeding an equal-selection benchmark (*p* < 0.001), indicating strong perceived near-term scalability in research services that the Dominican Republic’s clinical infrastructure is well positioned to support.

### Intellectual Property Protection and Institutional Coordination

Strengthening IP protection was viewed as a critical determinant of investment attractiveness: 89.3% of respondents assigned a high-importance rating to enhanced IP frameworks (binomial *p* < 0.001; Table A6). Complementing this, 73.8% of respondents expressed support for establishing a formal public–private biomedical coordination mechanism, with only 4.8% opposed (*p* < 0.001; Table A9). Taken together, these findings indicate that institutional trust and coordination capacity are perceived as foundational prerequisites—rather than secondary considerations—for scaling higher-value biomedical activity.

### Perceived Sector Competitiveness

Perceived competitiveness of the Dominican Republic’s biomedical sector relative to regional peers was moderate (mean = 3.24, SD = 0.98; Table A4). A one-sample *t*-test indicated that this mean did not differ significantly from the neutral midpoint (*t*(141) = 1.40, *p* = 0.163), characterizing an “emerging but not yet mature” competitive profile. Spearman’s rank correlation confirmed a significant positive association between perceived competitiveness and perceived government support (*r*s = 0.48, *p* < 0.001), consistent with the interpretation that visible policy execution and regulatory credibility are primary drivers of sector confidence.

### Workforce Skill Constraints

Reported talent shortages spanned the full biomedical value chain (Table A8). The most difficult profiles to source domestically were R&D scientists (81.0%), regulatory affairs specialists (78.6%), quality assurance/GMP engineers (73.8%), clinical research coordinators (71.4%), biostatisticians and data scientists (69.0%), and specialized manufacturing technicians (66.7%). Differences in reported shortage rates across roles were statistically significant (χ² = 15.2, *p* = 0.006), underscoring that both upstream scientific capacity and downstream regulatory-quality functions constitute binding constraints across the Triple Helix system.

### Priority Export and Collaboration Markets

Market priorities were strongly concentrated in the United States and North America (78.6%), with fewer endorsements for CARICOM and Caribbean markets (38.1%), South America (23.8%), and the European Union (16.7%; Table A10). A chi-square test confirmed that the distribution of market preferences differed significantly across options (χ² = 62.3, *p* < 0.001), reinforcing the manuscript’s emphasis on nearshoring and hemispheric partnership strategies as the primary short-to medium-term market development pathway.

### Multivariable Analyses

Exploratory multivariable models reinforced the centrality of governance quality and scientific capability as predictors of sector performance perceptions. In a linear regression model predicting perceived competitiveness, higher perceived government support (*β* = 0.42, *p* = 0.002), greater perceived talent availability (*β* = 0.31, *p* = 0.028), and higher confidence in IP protection (*β* = 0.29, *p* = 0.033) were each independently associated with greater perceived competitiveness, while perceived infrastructure adequacy did not reach statistical significance (*β* = 0.18, *p* = 0.21). Investor-respondent status was inversely but non-significantly associated with perceived competitiveness (β = −0.27, p = 0.09). The model explained 46% of the variance in competitiveness perceptions (*R²* = 0.46, *F*(5, 136) = 22.9, *p* < 0.001; Table A11).

A logistic regression model examining predictors of support for a formal public–private coordination body identified high regulatory concern (OR = 2.9, 95% CI [1.3, 6.4], *p* = 0.009), industry sector affiliation (OR = 2.4, 95% CI [1.1, 5.8], *p* = 0.021), and belief in the nearshoring opportunity (OR = 2.7, 95% CI [1.2, 6.1], *p* = 0.015) as significant positive predictors, whereas government sector affiliation was not significantly associated (OR = 0.71, 95% CI [0.3, 1.6], *p* = 0.41; Table A12). In a model of perceived foreign investment attractiveness, confidence in IP enforcement was the strongest predictor (*p* < 0.001), followed by perceived regulatory efficiency (*p* = 0.004) and workforce skill depth (*p* = 0.019; Table A13).

These exploratory models are based on a modest purposive sample, and their findings should be interpreted with appropriate caution. Nonetheless, they converge on a consistent conclusion: institutional credibility and advanced human capital are perceived as the principal levers for increasing sector competitiveness and attracting investment, a finding that maps directly onto the governance and university spheres of the Triple Helix framework.

### Executive Consultation: Thematic Synthesis

Qualitative analysis of executive consultation responses produced four convergent strategic themes that triangulate with and contextualize the quantitative survey findings.

### Embedded regulatory alignment as a structural asset

Executives emphasized that integration into U.S.-aligned supply chains has produced durable compliance capabilities—quality systems, traceability infrastructure, and industrial governance—that constitute genuine competitive assets rarely visible in aggregate trade statistics. This theme reinforces the survey finding that regulatory credibility is a primary driver of perceived competitiveness.

### Logistics speed as a nearshoring differentiator

Multiple respondents highlighted that ocean cargo can reach Miami in approximately 2.5 days, compared to approximately 40 days from Asia—an 86% reduction in lead time. Executives framed this not merely as a cost advantage but as a strategic enabler of “postponement” supply-chain strategies, whereby bulk imports are held for regional packaging and labelling in response to real-time demand signals.

### Clinical research as the primary value-chain upgrading pathway

Consistent with the survey’s finding that CRO services were the dominant sub-sector priority, executives identified clinical trials and evidence generation as the highest-potential near-term upgrading opportunity. The Dominican Republic’s combination of academic hospitals, patient population diversity, and competitive operational costs was seen as well-suited to regional clinical research hub development, contingent on regulatory harmonization with FDA standards.

#### Institutional fragmentation as the binding governance constraint

Across both government and industry respondents, the primary barrier was characterized not as an absence of capacity but as insufficient integration of existing capabilities—universities, regulators, hospitals, and trade agencies—into a coherent national strategy. This theme directly corroborates the survey’s statistical findings on academia–industry collaboration gaps and strong support for a formal coordination mechanism, and maps explicitly onto Triple Helix coordination failures documented in comparable middle-income innovation system contexts [8].

## Discussion

This study examined stakeholder perceptions of the Dominican Republic’s biomedical ecosystem through a concurrent mixed-methods design, integrating a cross-sectional survey of 142 domain experts with a strategic executive consultation. Three principal findings emerge. First, workforce shortages and academia–industry coordination failures are perceived as the binding constraints on sector upgrading, more so than market access, geography, or macroeconomic conditions. Second, regulatory credibility and IP enforcement are consistently identified as the highest-leverage institutional interventions, independently predicting perceived competitiveness, foreign investment attractiveness, and support for governance reform. Third, clinical trials and CRO services represent the most clearly perceived near-term pathway for value-chain upgrading, reflecting a convergent view among both survey respondents and executive consultants that the country’s clinical infrastructure is the most immediately scalable competitive asset.

Taken together, these findings are theoretically coherent with the Triple Helix model of innovation systems [8], which holds that knowledge-based economic upgrading requires the co-evolution of university, industry, and government roles and the quality of linkages among them. The Dominican Republic’s biomedical sector exhibits precisely the asymmetric helix configuration that the literature associates with stalled upgrading in middle-income economies: strong manufacturing and export performance within the industry sphere, but weak government–academia–industry coordination, underdeveloped research infrastructure, and regulatory institutions that have not yet matured to the standard required for higher-value activities [9,10].

### Governance and Regulatory Credibility

The finding that perceived government support is significantly below the neutral midpoint—and that it independently predicts both competitiveness and investment attractiveness—adds to a growing body of evidence that regulatory quality is not a secondary concern for biomedical sector development but a foundational one. Comparable findings have been documented in studies of Costa Rica’s life sciences sector, where CINDE’s sustained institutional engagement with multinational manufacturers—rather than wage or tax advantages alone—is credited as the primary driver of sectoral upgrading [11]. Ireland’s IDA similarly attributes its pharmaceutical sector leadership partly to predictable regulatory timelines and proactive alignment with European Medicines Agency standards [12].

In the Dominican Republic, the regulatory gap is compounded by the perceived absence of a formal coordination mechanism. The finding that 73.8% of respondents support a public–private biomedical authority—and that high regulatory concern is the strongest predictor of that support (OR = 2.9)—suggests that the demand for institutional reform is not rhetorical but actionable. WHO’s Good Regulatory Practices framework provides a directly applicable roadmap for this transition, including regulatory reliance models that allow smaller regulatory agencies to build credibility by referencing the decisions of stringent authorities such as the FDA and EMA [13]. Membership in the International Medical Device Regulators Forum (IMDRF), which the Dominican Republic recently joined with FDA support, provides an important foundational step. What remains is converting that membership into operational harmonization.

The salience of IP protection as a predictor of investment attractiveness—the strongest predictor in the foreign investment model—is consistent with the broader literature on pharmaceutical and biotech investment location decisions [14,15]. IP credibility functions less as a legal technicality than as a signal of institutional reliability: investors interpret strong IP enforcement as evidence that rule-of-law commitments are durable and that proprietary assets will be protected across political cycles. Strengthening IP frameworks is therefore as much a governance reform as a trade policy measure.

### Human Capital as the Primary Binding Constraint

Workforce shortages were the most frequently endorsed barrier (83.3%) and remained independently associated with competitiveness perceptions after controlling for governance and infrastructure variables. The pattern of shortages is analytically significant: gaps are reported not only in R&D scientists (81.0%) but also in regulatory affairs specialists (78.6%), quality assurance engineers (73.8%), biostatisticians (69.0%), and clinical research coordinators (71.4%). This distribution indicates that the constraint is not confined to the upstream innovation frontier but spans the full value chain, including the downstream compliance and quality functions that sustain manufacturing competitiveness.

This finding has direct implications for workforce strategy. Broad investments in science, technology, engineering, and mathematics (STEM) education, while necessary, are insufficient if they are not specifically calibrated to regulatory science, quality systems, and clinical operations—functions that are difficult to offshore or automate and that command sustained demand in both manufacturing and research service sectors. Singapore’s Economic Development Board has demonstrated that targeted sector-specific workforce programming, co-designed by government, universities, and industry, can compress the timeline for skill ecosystem development considerably [16]. The Dominican Republic’s diaspora embedded within the U.S. biomedical sector represents an additional, underutilised asset: diaspora engagement strategies have been shown to accelerate technology transfer, mentorship, and international network formation in comparable middle-income contexts [17].

### Clinical Trials and CRO Services as the Upgrading Pathway

The statistically dominant selection of clinical trials and CRO services as the priority sub-sector for expansion (76.2%, binomial *p* < 0.001) is consistent with broader global trends. IQVIA’s analysis of global clinical trial activity documents accelerating demand for trial sites in Latin America, driven by patient population diversity, competitive operational costs, and the availability of treatment-naïve patient populations in specific disease areas [18]. Clinical trials represent a qualitatively different form of value-chain participation than device assembly: they generate scientific knowledge, build domestic regulatory science capacity, create employment for highly trained professionals, and establish the international reputational credibility that attracts follow-on pharmaceutical investment.

The Dominican Republic’s combination of academic hospitals, geographic proximity to the North American market, and existing regulatory engagement with the FDA creates a credible foundation for CRO cluster development. Albert et al. [19] have argued that biomedical capacity in Caribbean states should be framed explicitly within a human security paradigm, positioning clinical research infrastructure not merely as an economic development asset but as a health-security imperative—one that reduces dependency on external diagnostic and therapeutic supply chains during crises. The COVID-19 pandemic made this dependency visible and costly [20], reinforcing the strategic case for domestic clinical research capacity as both an economic and a resilience investment.

### Hemispheric Supply-Chain Realignment and Nearshoring

The strong concentration of export market preferences toward the United States (78.6%) aligns the study’s findings with the broader post-pandemic reconfiguration of pharmaceutical and medical device supply chains. The National Academies of Sciences, Engineering, and Medicine [15] have documented the strategic vulnerability created by the concentration of pharmaceutical active ingredients and medical device manufacturing in Asia, and have recommended hemispheric diversification as a priority policy response. In this context, the Dominican Republic’s 2.5-day maritime transit time to Miami—compared with approximately 40 days from Asia—constitutes a structural competitive advantage that is not replicable through policy intervention alone.

The nearshoring frame also has implications for how the Dominican Republic positions its regulatory reform agenda. U.S. reshoring and friendshoring initiatives create demand not only for low-cost manufacturing capacity but also for manufacturing partners that can credibly meet FDA-equivalent quality standards, minimize the risk of supply disruptions, and operate in legally predictable environments. Regulatory harmonization with FDA standards is therefore not only a domestic governance priority but also a direct prerequisite for participation in the U.S. supply-chain resilience strategy—a point that the executive consultants in this study articulated clearly and that survey respondents’ prioritization of regulatory modernization (88.1%) corroborates.

### Comparison with Regional and International Benchmarks

The Dominican Republic’s current competitive position—moderate, not yet mature—can be usefully contextualized against two regional comparators and two international benchmarks. Costa Rica has achieved deep integration into U.S. medical device supply chains through a combination of free-zone infrastructure, targeted workforce development, and sustained government–industry partnership through CINDE; its sector generates over USD 3 billion in annual exports [11]. Puerto Rico demonstrates that FDA-aligned regulatory environments are sufficient to attract the highest-value pharmaceutical manufacturing, including biologics and sterile injectables [21]. Internationally, Singapore’s Biomedical Sciences Initiative illustrates the returns from deliberate, long-horizon government commitment to aligning research institutions, clinical infrastructure, and manufacturing in a single integrated ecosystem [16]. Ireland’s experience demonstrates that a small open economy can achieve global pharmaceutical leadership within two decades through institutional coherence, favorable tax conditions, and regulatory alignment [12].

What distinguishes the Dominican Republic from these comparators is not a deficit of foundational assets—its manufacturing base, trade access, and logistical advantages are comparable or superior—but the absence of the institutional coordination mechanism that in each comparator case served as the integrating architecture: CINDE in Costa Rica, the EDB in Singapore, and the IDA in Ireland. The study’s finding that 73.8% of stakeholders support establishing such a mechanism provides a clear and statistically robust mandate for this institutional step.

## Limitations

Several limitations should be noted. The survey relied on a purposive, non-probability sample, and results cannot be generalized to the full population of Dominican biomedical sector actors. Large-scale manufacturing operators appear underrepresented relative to their economic weight, which may have depressed responses related to logistics and infrastructure and amplified responses related to research and regulatory capacity. All measures are self-reported perceptions collected at a single point in time and may be subject to social desirability bias, sector advocacy effects, or recency influences from regulatory or market events occurring near the data-collection window.

The multivariable analyses are exploratory, and their findings should not be interpreted as causal estimates. The sample size of 142, while adequate for descriptive and bivariate inference, limits the power and stability of regression models with multiple covariates. Future research should seek to replicate these findings with larger and more representatively sampled populations, incorporate objective sector performance metrics alongside perceptual measures, and employ longitudinal designs capable of tracking changes in ecosystem conditions over time. Qualitative findings from the executive consultation, while systematically coded, reflect the perspectives of a small number of senior informants whose views may not be representative of broader leadership opinion.

## Conclusion

This study provides the first structured, mixed-methods assessment of stakeholder perceptions of the Dominican Republic’s biomedical ecosystem, integrating quantitative survey evidence from 142 domain experts with a qualitative executive consultation. The findings converge on a consistent and actionable conclusion: the Dominican Republic possesses genuine competitive foundations—manufacturing scale, logistical speed, trade access, and a growing human capital base—but has not yet developed the institutional architecture required to translate these assets into knowledge-intensive, higher-value biomedical participation.

The analysis, grounded in the Triple Helix model of innovation systems, identifies three structural priorities. First, regulatory modernization and IP credibility must be treated as foundational governance investments rather than sectoral administrative matters: they are the primary determinants of perceived competitiveness, foreign investment attractiveness, and confidence in the sector’s long-term trajectory. Second, workforce development must be redesigned around the specific skill profiles—regulatory science, quality assurance, clinical research operations, and data-intensive functions—that are both scarcest and highest-return in the current biomedical value chain. Third, the establishment of a formal biomedical public–private coordination authority, supported by an overwhelming majority of stakeholders and by comparative international evidence, would provide the institutional mechanism necessary to integrate existing assets into a coherent national strategy.

In a moment of significant hemispheric supply-chain reconfiguration, the Dominican Republic is strategically positioned to serve not merely as a manufacturing platform but as a trusted node for biomedical production, clinical research, and regional health security. Realizing this potential will require deliberate, sustained, and institutionally coherent policy action. The evidence presented here provides an empirical foundation for that agenda.

## Data Availability

NA

## Appendix A. Stakeholder survey instrument

**Table A1.** Dominican Republic biomedical industry stakeholder survey instrument.

| Item | Survey question | Response format / options |
| --- | --- | --- |
| 1 | Primary role or organization type within the Dominican Republic's biomedical ecosystem | Multiple choice (e.g., biotech/pharma, medical device manufacturing, research institution, government/regulator, healthcare provider, investor, service provider, other) |
| 1a | If 'Other', specify primary role | Short answer |
| 2 | Rate current level of government support for the biomedical industry in the Dominican Republic | Likert scale (1 = very poor; 5 = excellent) |
| 3 | Biggest challenges facing growth of the biomedical sector (select all that apply) | Multi-select (talent, funding, regulatory complexity, infrastructure, IP protection, market access, academia–industry collaboration, other) |
| 3a | If 'Other', specify additional challenge(s) | Short answer |
| 4 | Rate importance of the following factors for accelerating biomedical growth | Matrix Likert (not important to essential) covering R&D tax incentives; regulatory streamlining; specialized parks/free zones; international partnerships/technology transfer; access to skilled training |
| 5 | Sub-sectors with greatest promise for future growth (select up to three) | Multiple choice (clinical trials/CROs; medical devices; generic pharmaceuticals; digital health/health tech; biotechnology/biopharma; regenerative medicine) |
| 6 | Importance of enhancing IP protection for attracting foreign direct investment | Likert scale (1 = not important; 5 = essential) |
| 7 | Value of creating an organized biomedical industry public-private strategic entity/committee | Multiple choice (yes / no / maybe) |
| 8 | Optional email for receiving results or participating in follow-up | Short answer (optional) |
| 9 | Competitiveness of the Dominican biomedical sector compared with regional peers | Likert scale (1 = not competitive; 5 = highly competitive) |
| 10 | Most difficult professional skills to source domestically (select all that apply) | Multi-select (R&D scientists; regulatory specialists; quality assurance/GMP engineers; clinical research coordinators; biostatisticians/data scientists; specialized manufacturing technicians) |
| 11 | Priority international markets/regions for export and collaboration | Multiple choice (United States/North America; European Union; CARICOM/Caribbean; South America; Asia) |

## Appendix B. Supplementary survey results tables

**Table A2.** Respondent sector distribution (N = 142)

| Sector | n | % |
| --- | --- | --- |
| Research / Academic institutions | 44 | 31.0 |
| Healthcare providers | 34 | 23.8 |
| Biotech / Pharma / Medical devices | 24 | 16.7 |
| Government / Regulatory | 13 | 9.5 |
| Investors / Venture capital | 10 | 7.1 |
| CRO / Consulting / Services | 10 | 7.1 |
| Other | 7 | 4.8 |

**Table A3.** Perceived government support for biomedical sector (1–5 scale)

| Statistic | Value |
| --- | --- |
| Mean (SD) | 2.67 (1.12) |
| Median (IQR) | 3 (2–3) |
| Low (1–2) | 45.2% |
| Moderate (3) | 35.7% |
| High (4–5) | 19.1% |

Note: One-sample t-test $p = 0.034$ .

**Table A4.**
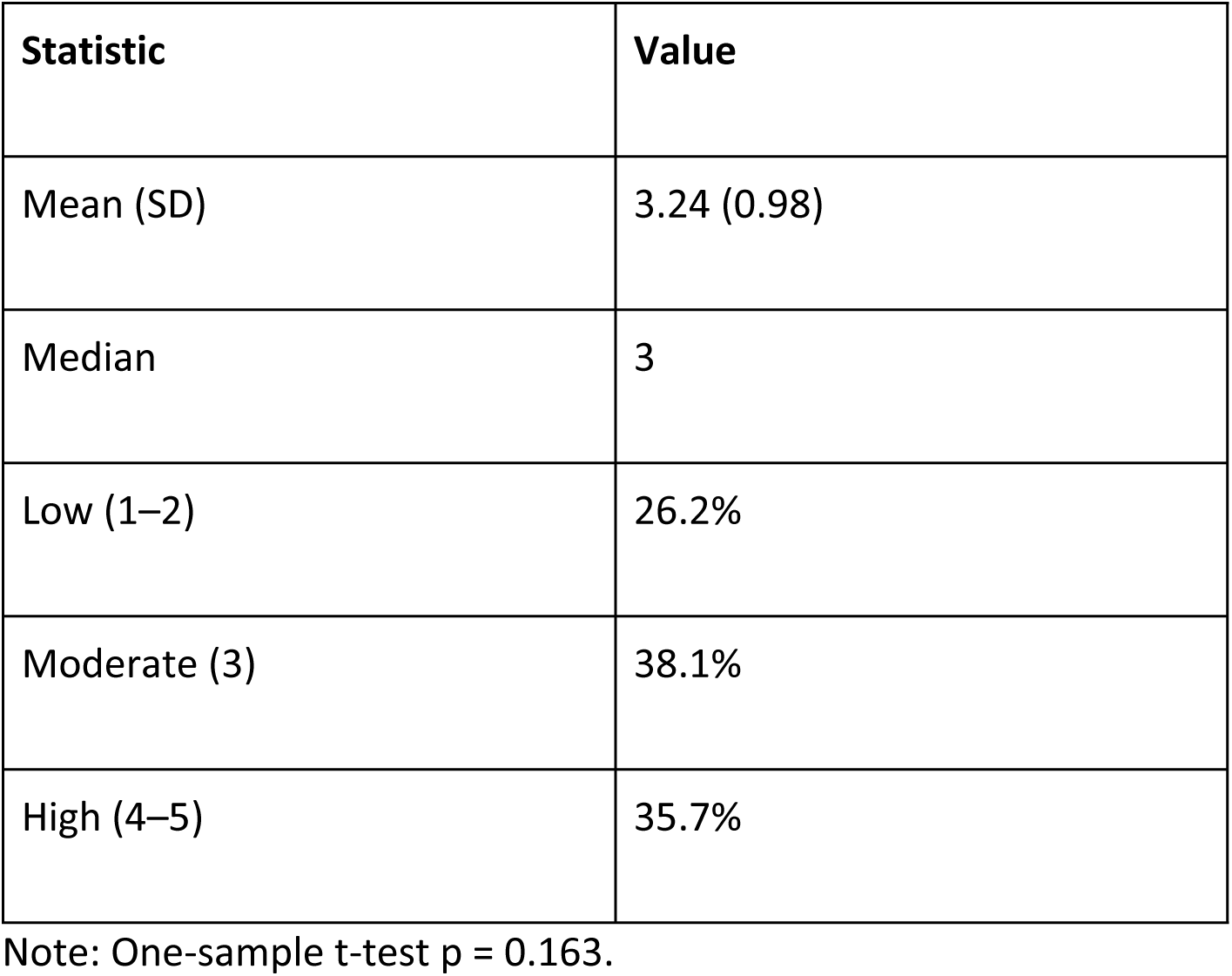
Perceived biomedical competitiveness relative to regional peers (1–5 scale)

| Statistic | Value |
| --- | --- |
| Mean (SD) | 3.24 (0.98) |
| Median | 3 |
| Low (1–2) | 26.2% |
| Moderate (3) | 38.1% |
| High (4–5) | 35.7% |

Note: One-sample t-test $p = 0.163$ .

**Table A5.** Major structural barriers to biomedical sector growth (multi-select)

| Barrier | % selecting |
| --- | --- |
| Specialized talent shortages | 83.3% |
| Insufficient funding / capital | 76.2% |
| Weak academia–industry collaboration | 71.4% |
| Infrastructure gaps | 69.0% |
| Regulatory delays / complexity | 66.7% |
| IP protection concerns | 61.9% |

Note: Chi-square test for non-uniform clustering across barrier categories: $\chi^2(5) = 18.7$ , $p = 0.002$ .

**Table A6.** High-importance ratings (very important/essential) for priority levers and IP protection.

| Policy lever / enabling factor | % high importance |
| --- | --- |
| Regulatory modernization | 88.1% |
| Workforce development | 85.7% |
| International partnerships and technology transfer | 83.3% |
| R&D tax incentives | 81.0% |
| Biomedical free-zones / specialized parks | 78.6% |
| IP protection (importance for attracting foreign direct investment) | 89.3% |

Note: Among the policy lever items, the Friedman test indicated differences in relative priority ranks ( $p = 0.005$ ). For the IP protection item, 89.3% rated high importance (binomial $p < 0.001$ ).

**Table A7.**
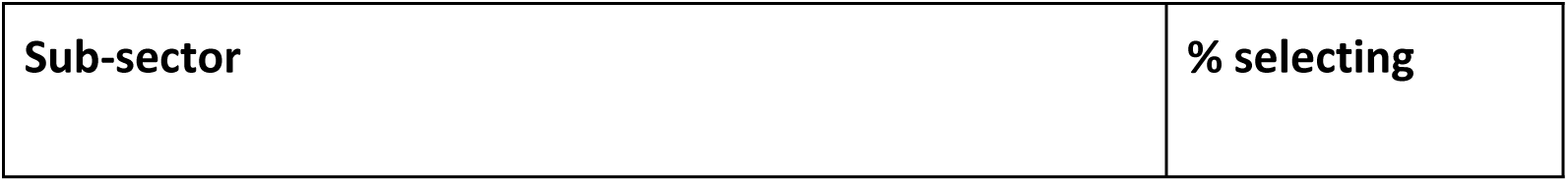
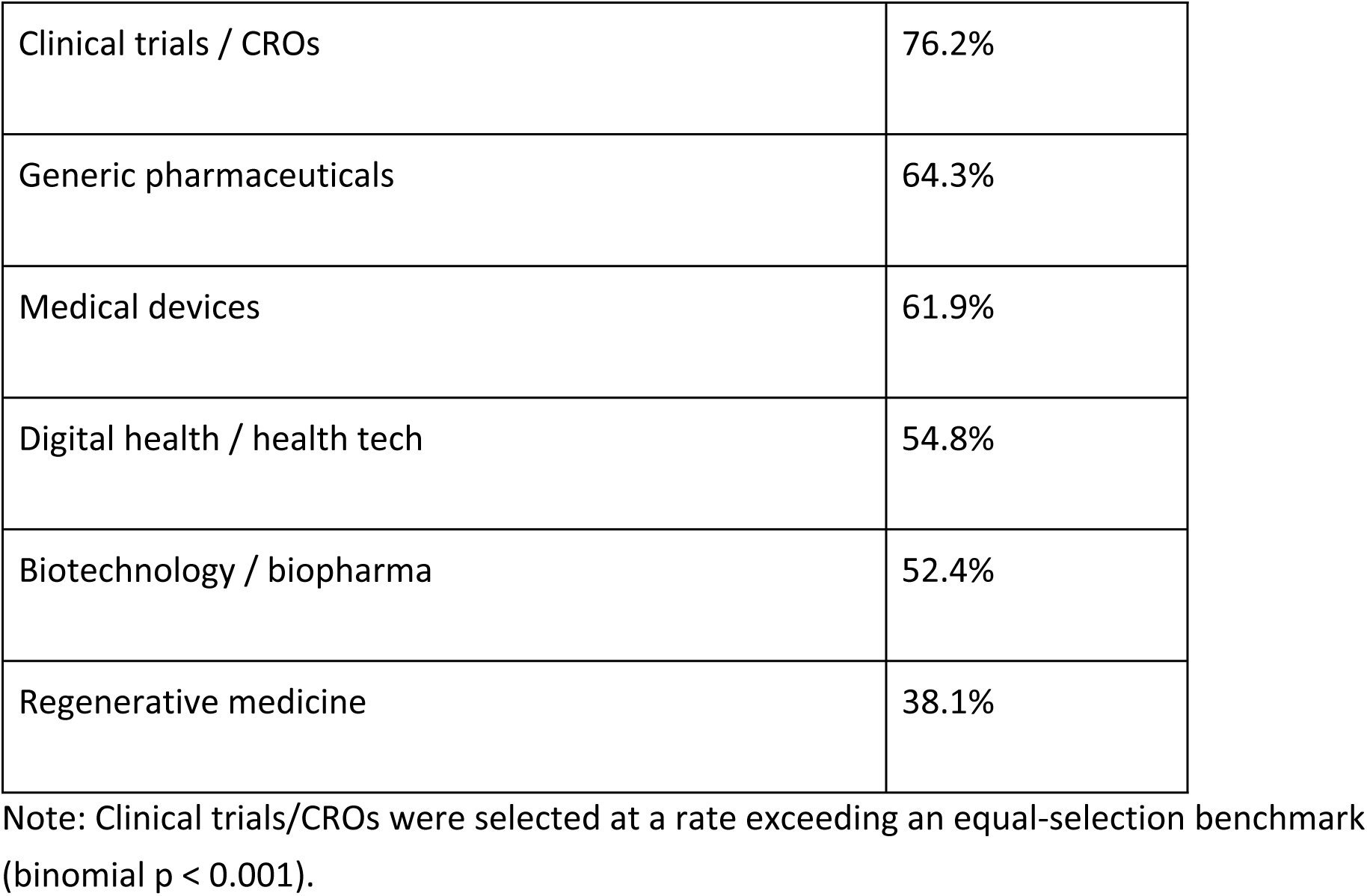
Biomedical sub-sectors with greatest perceived growth potential (select up to three)

**Table A8.**
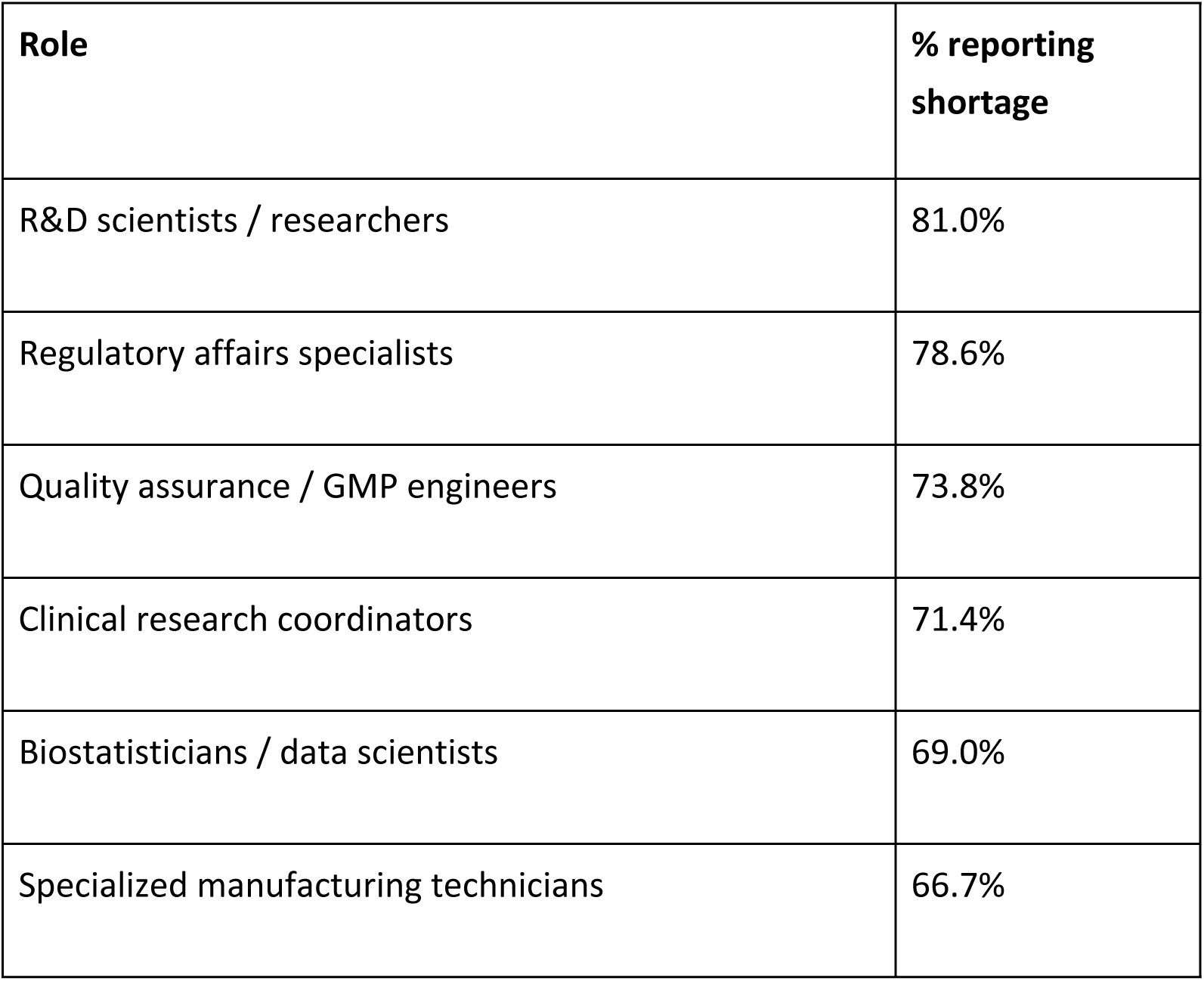
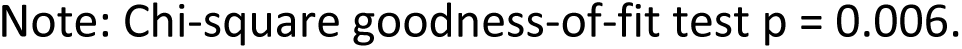
Workforce skills most difficult to source domestically (multi-select)

| **Role** | **% reporting shortage** |
| --- | --- |
| R&D scientists / researchers | 81.0% |
| Regulatory affairs specialists | 78.6% |
| Quality assurance / GMP engineers | 73.8% |
| Clinical research coordinators | 71.4% |
| Biostatisticians / data scientists | 69.0% |
| Specialized manufacturing technicians | 66.7% |

Note: Chi-square goodness-of-fit test $p = 0.006$ .

**Table A9.** Support for a public–private biomedical governance entity.

| Response | % |
| --- | --- |
| Yes | 73.8% |
| Maybe / unsure | 21.4% |
| No | 4.8% |

Note: Chi-square test $p < 0.001$ .

**Table A10.** Priority markets for export and collaboration.

| Market / region | % selecting |
| --- | --- |
| United States / North America | 78.6% |
| CARICOM / Caribbean | 38.1% |
| South America | 23.8% |
| European Union | 16.7% |

Note: Chi-square test comparing market selection $p < 0.001$ .

**Table A11.** Linear regression predictors of perceived sector competitiveness.

| Predictor | $\beta$ | SE | p |
| --- | --- | --- | --- |
| Perceived government support | +0.42 | 0.12 | 0.002 |
| Talent availability | +0.31 | 0.14 | 0.028 |
| Confidence in IP protection | +0.29 | 0.13 | 0.033 |
| Infrastructure adequacy | +0.18 | 0.15 | 0.21 |
| Investor respondent (vs others) | −0.27 | 0.16 | 0.09 |

Note: Model $R^2 = 0.46$ ; overall model $p < 0.001$ .

**Table A12.** Logistic regression predictors of support for a public–private governance body.

| Predictor | OR | 95% CI | p |
| --- | --- | --- | --- |
| High regulatory concern | 2.9 | 1.3–6.4 | 0.009 |
| Industry respondent | 2.4 | 1.1–5.8 | 0.021 |
| Belief in nearshoring opportunity | 2.7 | 1.2–6.1 | 0.015 |
| Government respondent | 0.71 | 0.3–1.6 | 0.41 |

**Table A13.** Predictors of perceived foreign investment attractiveness (linear model)

| Predictor | $\beta$ | p |
| --- | --- | --- |
| Confidence in IP enforcement | +0.51 | <0.001 |
| Regulatory efficiency | +0.37 | 0.004 |
| Workforce skill depth | +0.28 | 0.019 |

---

**Figure 1.**
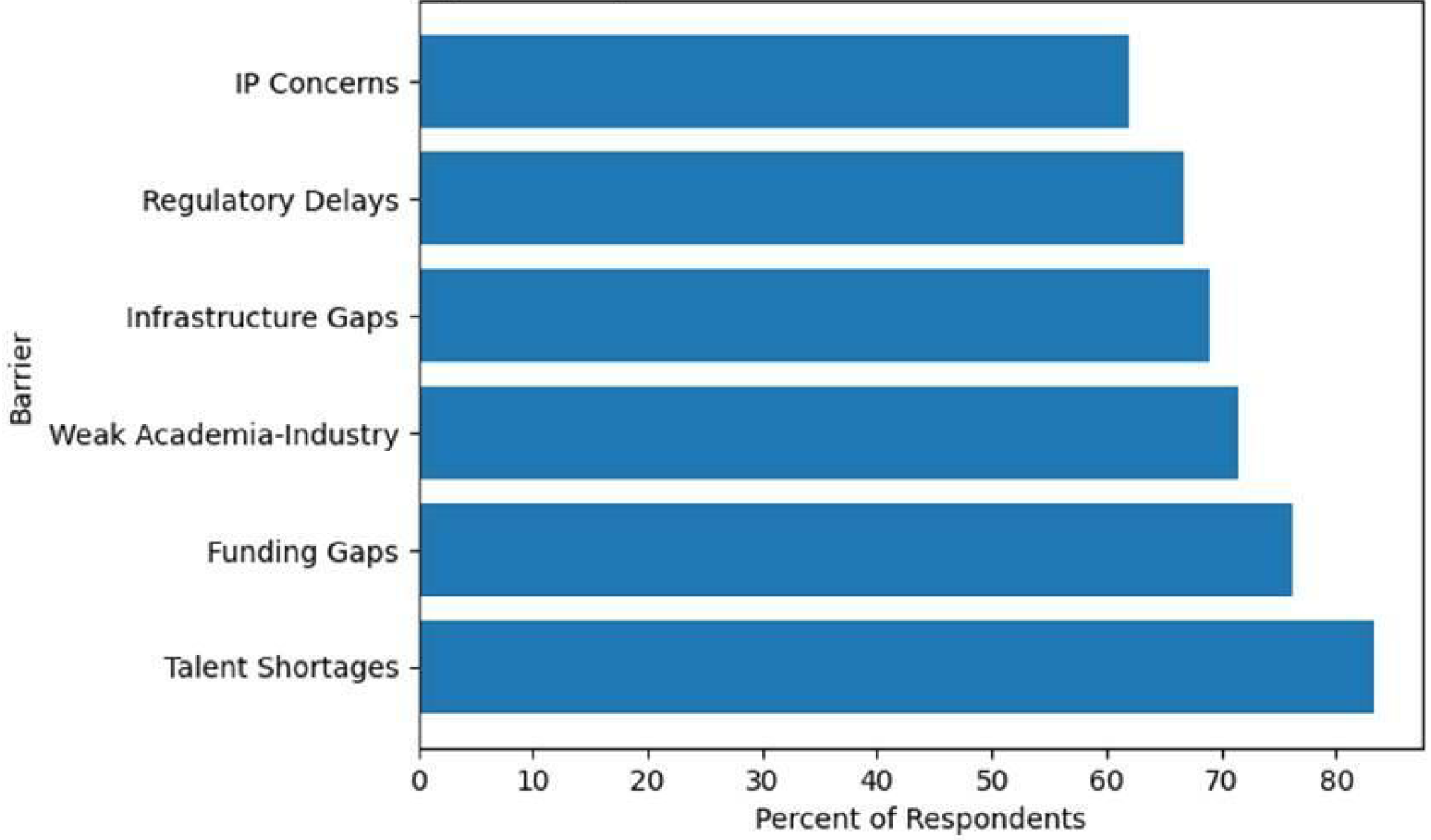
Major Structural Barriers to Biomedical Sector Growth. Barriers were not evenly distributed across categories (χ²(5) = 18.7, p = 0.002). Talent shortages (83.3%) and funding gaps (76.2%) were the most frequently cited constraints, followed by weak academia–industry collaboration (71.4%), infrastructure gaps (69.0%), regulatory delays (66.7%), and intellectual property concerns (61.9%).

**Figure 2.**
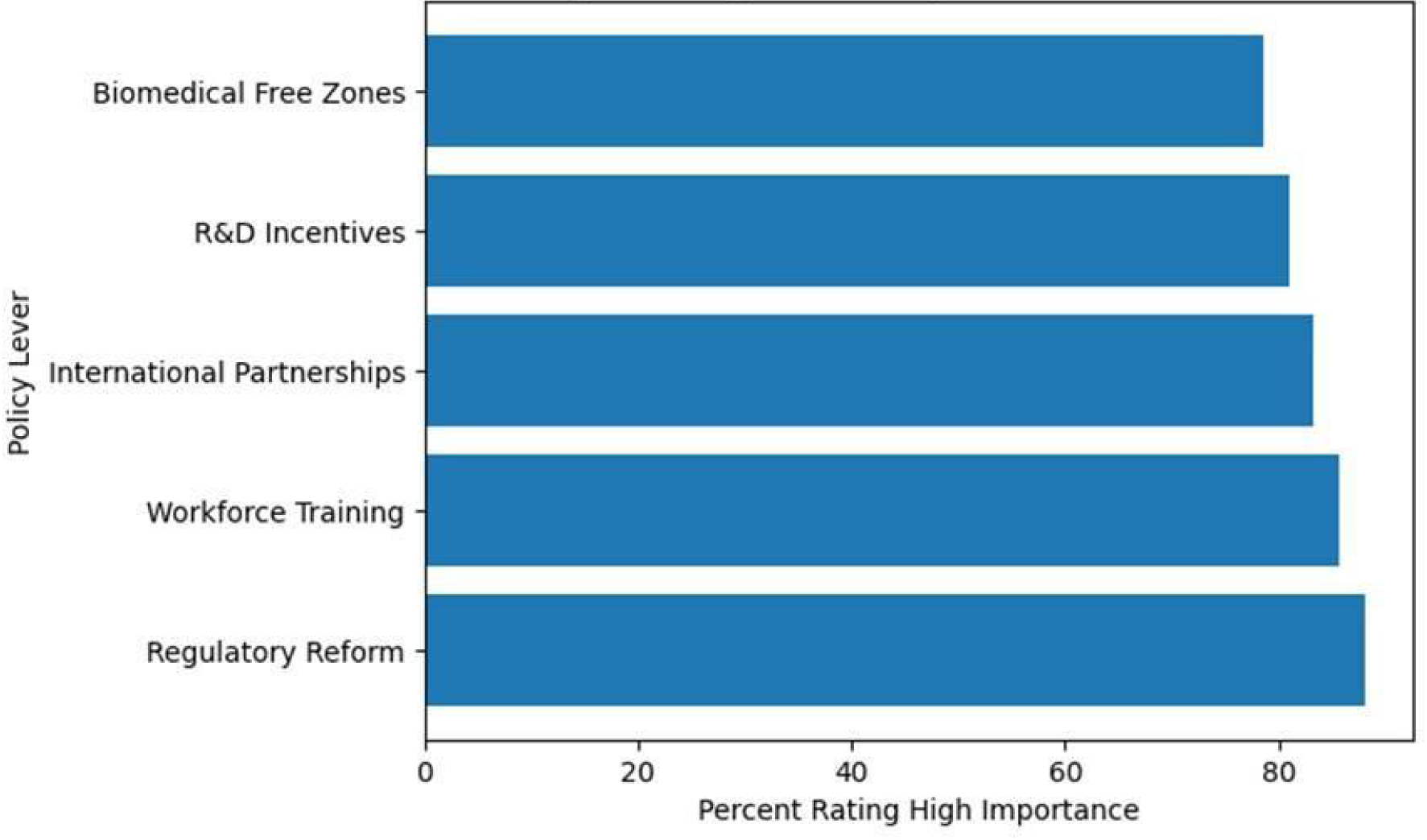
Highest-Priority Policy Levers for Biomedical Growth. Regulatory reform (88.1%) and workforce development (85.7%) ranked highest among respondents. Priority rankings differed significantly across policy levers (Friedman χ²(4) = 14.9, p = 0.005), indicating strong consensus around governance and human-capital interventions.

**Figure 3.**
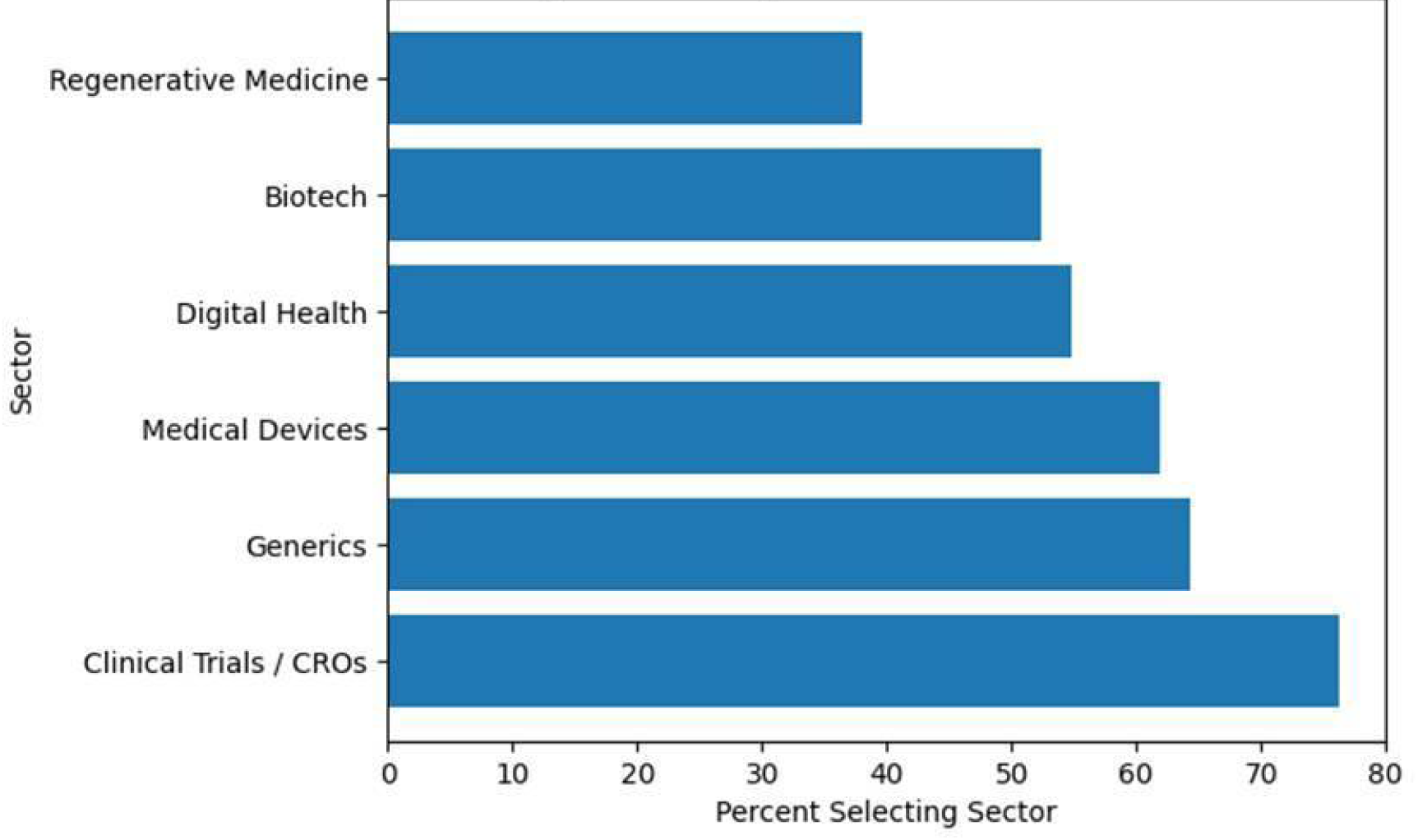
Biomedical Sub-Sectors with the Greatest Perceived Growth Potential. Clinical trials and CRO services were the most frequently selected opportunity (76.2%) and were statistically dominant relative to other sectors (binomial test, p < 0.001). Generics (64.3%), medical devices (61.9%), and digital health (54.8%) represent additional near-term expansion opportunities.

**Figure 4.**
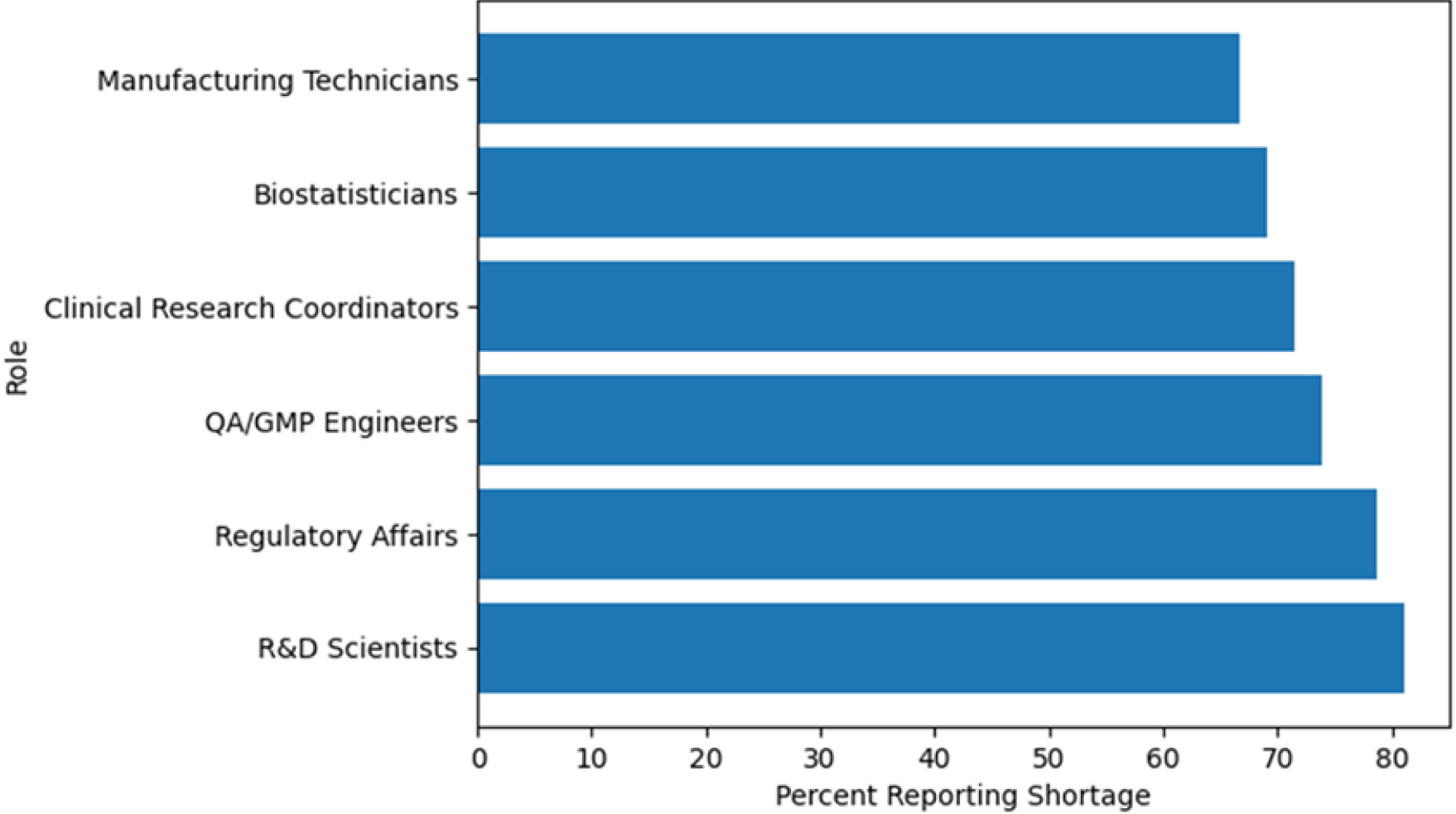
Biomedical Workforce Talent Shortages. Shortages were most pronounced in R&D scientists (81.0%), regulatory affairs (78.6%), and QA/GMP engineering (73.8%). Role-specific shortages differed significantly p = 0.006), confirming workforce capacity as a binding constraint.

**Figure 5.**
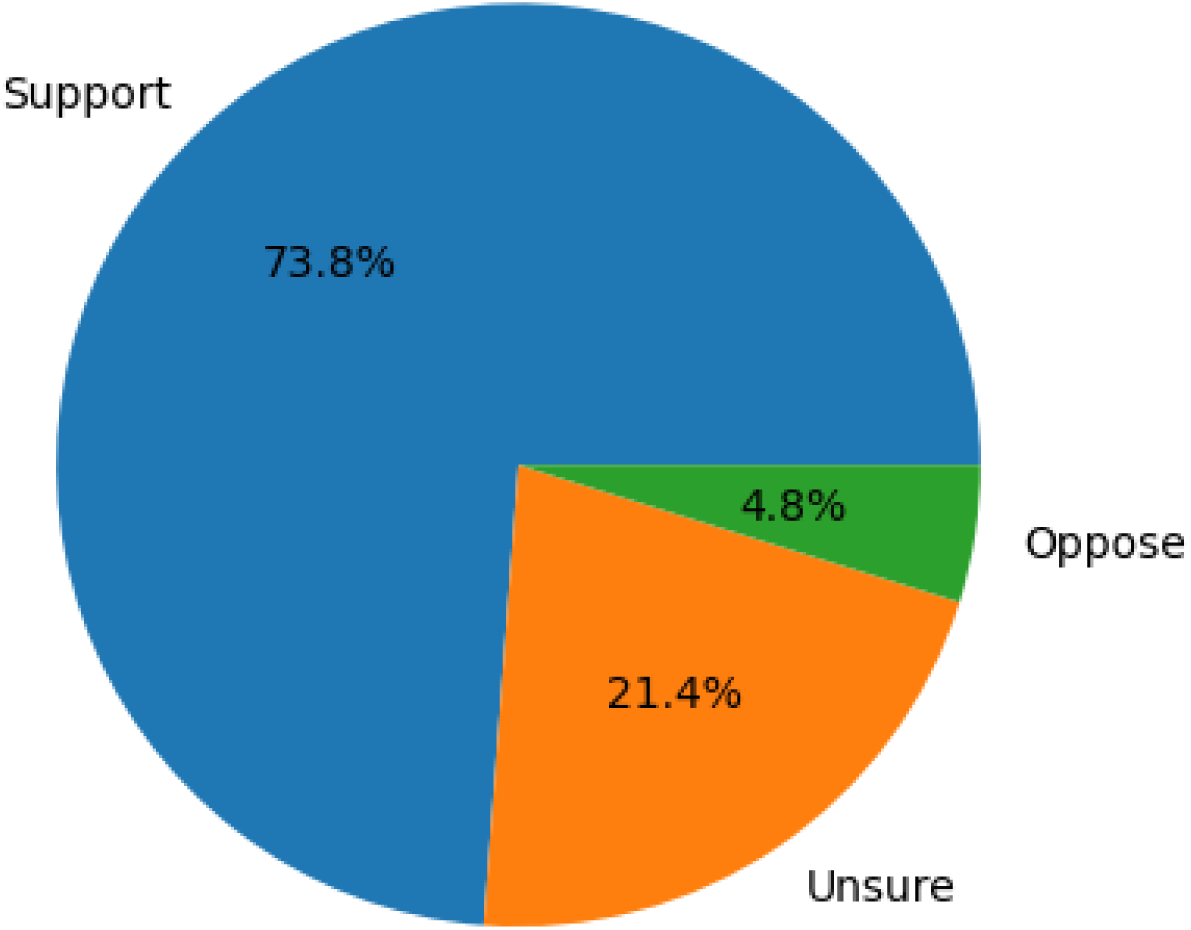
Stakeholder Support for a Public-Private Biomedical Governance Entity. A strong majority supported the creation of a national biomedical coordinating body (73.8%), with only 4.8% opposed (p < 0.001), indicating ecosystem readiness for structured governance.

## Appendix C. Executive consultation protocol

Senior leaders from the public sector and industry were invited to provide strategic perspectives on the role of the Dominican biomedical sector in hemispheric health security. Participants were asked to respond to two prompts: (1) What unique strengths does the Dominican Republic possess to become a pillar of the biomedical industry and contribute to hemispheric health security? and (2) What concrete opportunities do you see for the Dominican biomedical industry to strengthen health resilience and strategic autonomy across the Americas over the next five years? Responses were submitted either in written form or as audio recordings and were synthesized thematically to inform the narrative in the Executive Consultation: Thematic Synthesis section.

